# Multi-omics highlights ABO plasma protein as a causal risk factor for COVID-19

**DOI:** 10.1101/2020.10.05.20207118

**Authors:** Ana I. Hernández Cordero, Xuan Li, Stephen Milne, Chen Xi Yang, Yohan Bossé, Philippe Joubert, Wim Timens, Maarten van den Berge, David Nickle, Ke Hao, Don D. Sin

## Abstract

SARS-CoV-2 is responsible for the coronavirus disease 2019 (COVID-19) and the current health crisis. Despite intensive research efforts, the genes and pathways that contribute to COVID-19 remain poorly understood. We therefore used an integrative genomics (IG) approach to identify candidate genes responsible for COVID-19 and its severity. We used Bayesian colocalization (COLOC) and summary-based Mendelian randomization to combine gene expression quantitative trait loci (eQTLs) from the Lung eQTL (n=1,038) and eQTLGen (n=31,784) studies with published COVID-19 genome-wide association study (GWAS) data from the COVID-19 Host Genetics Initiative. Additionally, we used COLOC to integrate plasma protein quantitative trait loci (pQTL) from the INTERVAL study (n=3,301) with COVID-19-associated loci. Finally, we determined any causal associations between plasma proteins and COVID-19 using multi-variable two-sample Mendelian randomization (MR). We found that the expression of 20 genes in lung and 31 genes in blood was associated with COVID-19. Of these genes, only three (*LZTFL1, SLC6A20* and *ABO*) had been previously linked with COVID-19 in GWAS. The novel loci included genes involved in interferon pathways (*IL10RB, IFNAR2* and *OAS1*). Plasma ABO protein, which is associated with blood type in humans, demonstrated a significant causal relationship with COVID-19 in MR analysis; increased plasma levels were associated with an increased risk of having COVID-19 and risk of severe COVID-19. In summary, our study identified genes associated with COVID-19 that may be prioritized for future investigation. Importantly, this is the first study to demonstrate a causal association between plasma ABO protein and COVID-19.

## Introduction

The severe acute respiratory syndrome coronavirus 2 (SARS-CoV-2) is responsible for the coronavirus disease 2019 (COVID-19) and the current world pandemic. The SARS-CoV-2 was first identified in Wuhan, Hubei province, China in late 2019 and since then, it has spread across the world, affecting more than 180 countries (Zhu et al. 2020). To date, the COVID-19 health crisis has resulted in the loss of 973,443 human lives (Dong et al. 2020). While the disease has mild effects in most individuals, severe COVID-19 is more likely in the elderly population and individuals with comorbidities such as cardiovascular diseases and diabetes (Zhou et al. 2020a). Why these populations are at a higher risk of adverse COVID-19 outcomes is still unclear.

Genetic variations in the host may explain some of the heterogeneity in COVID-19 outcomes. Understanding the role of genetic variants may also provide critical insights into COVID-19 pathogenesis. However, the genes and pathways that contribute to SARS-CoV-2 infection are poorly understood. Recently, a pooled genome-wide association study (GWAS) revealed novel susceptibility loci, including 3p21.31 and 9q34.2, for severe COVID-19 related to respiratory failure (Ellinghaus et al. 2020). These loci encompassed several genes, but which (if any) of the individual genes are causally associated with COVID-19 was not explored.

GWAS leverages genetic variants to determine associations between regions of the genome and a particular trait (e.g. disease). However, traditional GWAS has several limitations that may prevent true gene-trait associations from being identified. For example, due to correction for multiple comparisons, GWAS results necessarily have a stringent threshold for statistical significance and relevant associations below this threshold may be missed. Second, disease-associated loci are typically thousands of base pairs wide and thus contain multiple genes, which may obscure the causal gene contributing to the disease trait. One emerging approach to identify genes within susceptibility loci is integrative genomics. By combining genomic information with transcriptomic, proteomic and/or methylation data, integrative genomics is able to fine-map genetic susceptibility loci and identify genes and proteins most likely to have a causal association with disease. This method has previously elucidated specific protein coding genes and mechanisms that contribute to complex traits (Cano-Gamez and Trynka 2020). In the present study, we harnessed the power of integrative genomics to identify several susceptibility genes for COVID-19 and investigate the causal relationship between the plasma protein levels of candidate genes and COVID-19. Notably, here, we demonstrate a causal association of the ABO protein (this protein is responsible for the ABO blood groups) with both the risk for COVID-19 and its severity.

## Methods

### Study cohorts

#### COVID-19 Host Genetics Initiative (COVID-19 HG)

For our study we obtained publicly available summary statists from the COVID-19 HG GWAS meta-analysis V3 (https://www.covid19hg.org/) (The COVID-19 Host Genetics Initiative 2020). We obtained the summary statistics for two case-control analyses: 1) “susceptibility to COVID-19” (where cases were all individuals with a diagnosis of COVID-19 [n=6,696], and controls were all individuals without a COVID-19 diagnosis [n=1,073,072]); and 2) “Severe COVID-19” (where cases were individuals with a COVID-19 diagnosis and who were hospitalized [n = 3,199], and controls were all individuals without a COVID-19 diagnosis and no hospitalization [n = 897,488]). In brief, the COVID-19 HG is an ongoing initiative that aims to shared resources and facilitate COVID-19 genetic host research through international collaboration. Contributing investigators submit individual level data or summary results from GWASs performed according to pre-specified methodological standards to ensure quality and consistency of the data. For imputation each individual study used their own reference panel or existing imputation panel; the association analyses were adjusted for age, sex and population structure. The COVID-19 HG combines this data in an inverse variance weighted meta-analysis using variants with minor allele frequency (MAF) > 0.0001 and imputation quality (r^2^) > 0.6. Further details are provided by the COVID-19 HG (The COVID-19 Host Genetics Initiative 2020).

#### Lung eQTL study

For our study we obtained lung expression quantitative loci (eQTL) from the eQTL study. In summary, the lung eQTL study cohort consisted of 1,038 participants from three institutions, University of British Columbia (UBC), Laval University and University of Groningen. At UBC and Laval University the studies were approved by the ethics committees within each institution. For University of Groningen the lung specimens were provided by the local tissue bank of the Department of Pathology, the study protocol used was consistent with the Research Code of the University Medical Center Groningen and Dutch national ethical and professional guidelines (“Code of conduct; Dutch federation of biomedical scientific societies”, http://www.federa.org). This study determined the gene expression of non-tumour lung tissue samples using 43,466 non-control probe sets (see GEO platform GPL10379). The participants were also genotyped using the Illumina Human 1M Duo BeadChip, and after imputation a total of 7,640,142 single nucleotide polymorphisms (SNPs) for Laval, 7,610,179 for UBC, and 7,741,505 for Groningen were kept for an eQTL analysis. Data for each site were evaluated, separately, using a linear regression model, which assumed an additive genotype effect. Site-specific results were then combined by meta-analysis using a fixed effects model with inverse variance weighting. cis expression quantitative trait loci (cis-eQTLs) were defined by a 2 Mb window (± 1Mb probe to SNP distance). Full details of the cohort and genotyping quality control, and eQTL analysis are provided by Hao and colleagues (Hao et al. 2012).

#### eQTLGen

We obtained blood cis-eQTL summary statistics from eQTLGen. The eQTLGen cohort used to estimate the cis-eQTLs consisted of 31,684 whole blood (85%) and peripheral blood mononuclear cell (15%) samples from 37 datasets. Gene expression profiles and genotypes were obtained for the eQTLGen cohort. The full details on the participants, gene expression processing and genotyping for each dataset are described by the eQTLGen consortium (Võsa et al. 2018). The cis-eQTL analysis was performed in each separate dataset and were estimated within a 2 Mb window (± 1Mb probe to SNP distance) as previously described by Westra and colleagues (Westra et al. 2013), later the results were combine by meta-analysis using a weighted Z-score method (Westra et al. 2013; Võsa et al. 2018).

#### INTERVAL study

We used plasma protein quantitative trait loci (pQTL) obtained from the INTERVAL study (Sun et al. 2018). In brief, the INTERVAL study was a randomized trial of blood donation intervals that comprises around 50,000 participants from 25 static donor centres of NHS Blood and Transplant (NHSBT) (Di Angelantonio et al. 2017; Sun et al. 2018). Blood was collected from the participants using standard venepuncture. The Affymetrix Axiom UK Biobank array was used for the genotyping of 830,000 SNPs and genotypes were later imputed using the 1000 Genome phase 3 UK10K reference panel. After quality controls 10,572,788 SNPs were retained. A randomly-selected subset of 3,301 participants were used for the plasma pQTL analyses of 3,622 proteins (Sun et al. 2018). Plasma protein levels were measured by using an expanded version of an aptamer-based multiplex protein assay (SOMAscan) previously described by Sun and colleagues(Sun et al. 2018). The protein levels were adjusted for confounding variables (age, sex, waiting period between blood collection and processing and the first three genetic principal components) and the residuals were extracted and rank-inverse normalized. pQTL analysis consistent of testing genetic associations with a linear regression using an additive genetic model. The results from each donor centers were combined using fixed-effect inverse-variance meta-analysis. Further details on the study cohort, genotyping protocol and quality control are described by Sun and colleagues (Sun et al. 2018).

### Integrative-omics methods

#### Bayesian Colocalization test (Coloc)

We first conducted Coloc tests to determine the probability that SNPs associated to COVID-19 phenotypes and gene expression (eQTLs) were consistent with shared genetic causal variants (colocalization). This integrative genomic method estimates the ‘posterior probabilities’ (PP) of five hypothesis described as follow: a genetic locus has no associations with either of the two traits (i.e. gene expression and a complex trait) investigated (H_0_); the locus is associated only with gene expression (H_1_); the locus is associated only with the complex trait (H_2_); the locus is associated with both traits via independent SNPs (H_3_); the locus is associated with both traits through shared SNPs (H_4_) (i.e.: a SNP is associated with COVID-19 and is also a cis-eQTL). Colocalization is therefore indicated by a high PP of H_4_ being true. For these analyses we used the coloc package(Giambartolomei et al. 2014) implemented in R. We tested only cis-eQTL regions (± 1Mb probe to SNP distance). As required by the method and recommended by Giambartolomei and colleagues (Giambartolomei et al. 2014) we set ‘prior probability’ of the various configurations (H_1,_ H_2,_ and H_4_). For the eQTL dataset we used 1 × 10^−04^ prior probability for a cis-eQTL (H_1_). We also used 1 × 10^−04^ prior probability for COVID-19 associations (H_2_). Finally, we set a prior probability that a single variant affects both traits (H_4_) to be 1 × 10^−06^. We set significant colocalization (posterior probability) at *PP*_*H4*_ > 0.80. We executed coloc between the loci associated to COVID-19 (susceptibility and disease severity) and cis-eQTLs associated to gene expression in both lung and blood tissues, retaining genes whose expression colocalized with COVID-19 in both compartments as ‘candidate genes’. If the corresponding proteins of the candidate genes were present in the plasma protein dataset (INTERVAL study) we executed Coloc analysis for the plasma protein levels and COVID-19 phenotypes.

#### Summary-based Mendelian Randomization (SMR)

The SMR method was specifically built to test the association between gene expression and a complex trait using a SNP as the instrument (Zhu et al. 2016). SMR is based on the standard Mendelian Randomization (MR) analysis, where the effect of genetic variants are linked to the trait of interest via an exposure (gene expression) (Swerdlow et al. 2016). Therefore, we employed SMR method to identify genes whose expression in lung and blood may mediate the effect of genetic variants on COVID-19. For the SMR the lung and blood cis-eQTLs and COVID-19 HG GWAS meta-analysis summary statistics were used. We selected the 1000G phase 3 EUR (1000 Genomes Project Consortium et al. 2015) as the reference panel for linkage disequilibrium (LD) estimation. Significant SMR was defined at *P*_*SMR*_ < 0.001. The significant SMR by itself does not necessarily means that the same variants are associated with the gene expression and the phenotype; the association could be a consequence of the LD between independent causal variants, rather than pleiotropy of a single causal variant or causality. To determine whether the associations were related to LD we used the heterogeneity in dependent instruments test (HEIDI) test (Zhu et al. 2016), the null hypothesis of which is that the effect of a variant is shared for gene expression and the phenotype; rejecting the null hypothesis based on the p value (*P*_*HEIDI*_) is interpreted as evidence of heterogeneity (linkage) (Zhu et al. 2016). We reported the significant SMR associations that also show *P*_*HEID*_ > 0.05.

#### Mendelian Randomization: ABO plasma protein and COVID1-9

To test causality of the ABO plasma protein we conducted MR tests of the plasma protein on COVID-19 phenotypes. MR is based on the unidirectional flow of genetic information, which assumes that genetic variants will have an effect on downstream phenotypes (gene protein phenotype). MR uses SNPs as instrumental variables (IVs) to link a risk factor (‘exposure’) to a health trait (‘outcome’). The MR assumptions are as follow: IVs are associated with the exposure, and only affect an outcome *via* the exposure, and are independent of confounders.

A pQTL on chromosome 9q34.2 was identified for ABO plasma protein (Sun et al. 2018). We performed stepwise conditional analysis, using GCTA 1.92.0beta3 (Yang et al. 2011), within each 2 Mb region on chromosome 9 to identify independently associated SNPs. UK Biobank genotypes (Bycroft et al. 2018) were used as the reference sample, and excluded SNPs with MAF < 0.01. We later extracted the effect size (beta) and standard error (SE) for each independent variant. Likewise, we obtained the beta and SE for each of these SNPs on the two COVID-19 phenotypes in the COVID-19 HG meta-analysis. We then performed an inverse variance weighted (IVW) MR (IVW-MR) (implemented in ‘MendelianRandomization’ R package (Staley 2020)) to link the allelic effects on the exposure (plasma protein levels) to their effects on the outcome (severe COVID-19 and COVID-19 diagnosis). We adjusted for possible correlations between SNPs by incorporating the estimated LD on 1000G phase 3 EUR using PLINK 1.9 (Chang et al. 2015) into MR input. The IVW-MR assumes no directional pleiotropy (i.e. genetic variant associated with multiple unrelated phenotypes) (Bowden et al. 2015). Significance of the MR estimate was set at *P* < 0.05. To assess the presence of pleiotropy we then executed MR-Egger analysis (Bowden et al. 2015) which, unlike IVW-MR, allows the estimation of directional pleiotropy. The presence of pleiotropy is suggested by a significant (*P* _*Egger Intercept*_ < 0.05), non-zero, intercept term.

We assessed heterogeneity of the IVs using the Cochran’s Q test obtained from the IVW-MR output (Staley 2020); significant heterogeneity (*P* < 0.05) suggests that the variability of the IVs estimates is greater than would be expected by chance alone, which may indicate bias due to invalid IVs. Although a MR with multiple IVs has greater power compared to a single IV MR, it is possible that the average causal effect may be mainly driven by the SNP with the strongest association with the outcome. To evaluate the reliance of the multi-variable MR on individual SNPs we performed a sensitivity analysis by excluding each SNP at a time and re-calculating the IVW-MR estimate. If removing a SNP significantly changes the MR estimate, it indicates that the multi-variable MR relied mostly on the removed SNP.

## Results

### Integration of gene expression and COVID-19 genomics reveals novel gene associations

The COVID-19 HG GWAS identified one genetic locus associated with susceptibility to COVID-19 (defined as a positive COVID-19 diagnosis versus the general population) and one genetic locus associated with severe COVID-19 (defined as hospitalization for COVID-19 versus the general population) at a genome-wide significant threshold of *P*_*GWAS*_ < 5 × 10^−08^. In our analysis, in addition to these loci, we also explored regions below this stringent threshold. We integrated these GWAS results with gene expression data from both lung tissue (Lung eQTL Study) and blood (eQTLGen) using two statistical methodologies (described in detail in the Methods) (**Fig. 1**). Bayesian colocalization assess whether two genetic association signals are consistent with a shared causal variant. We defined colocalization as the posterior probability of this hypothesis (*PP*_*H4*_) being > 0.80. SMR integrates summary data from GWAS and eQTLs in order to identify genes whose expression levels are associated with a trait due to the effects of a common genetic variant (either by direct causal or pleiotropic effects) rather than due to genetic linkage. Significance of SMR estimate was set at *P*_*SMR*_ < 0.001 with no significant heterogeneity (*P*_*HEIDI*_ > 0.05).

**Fig. 1.**
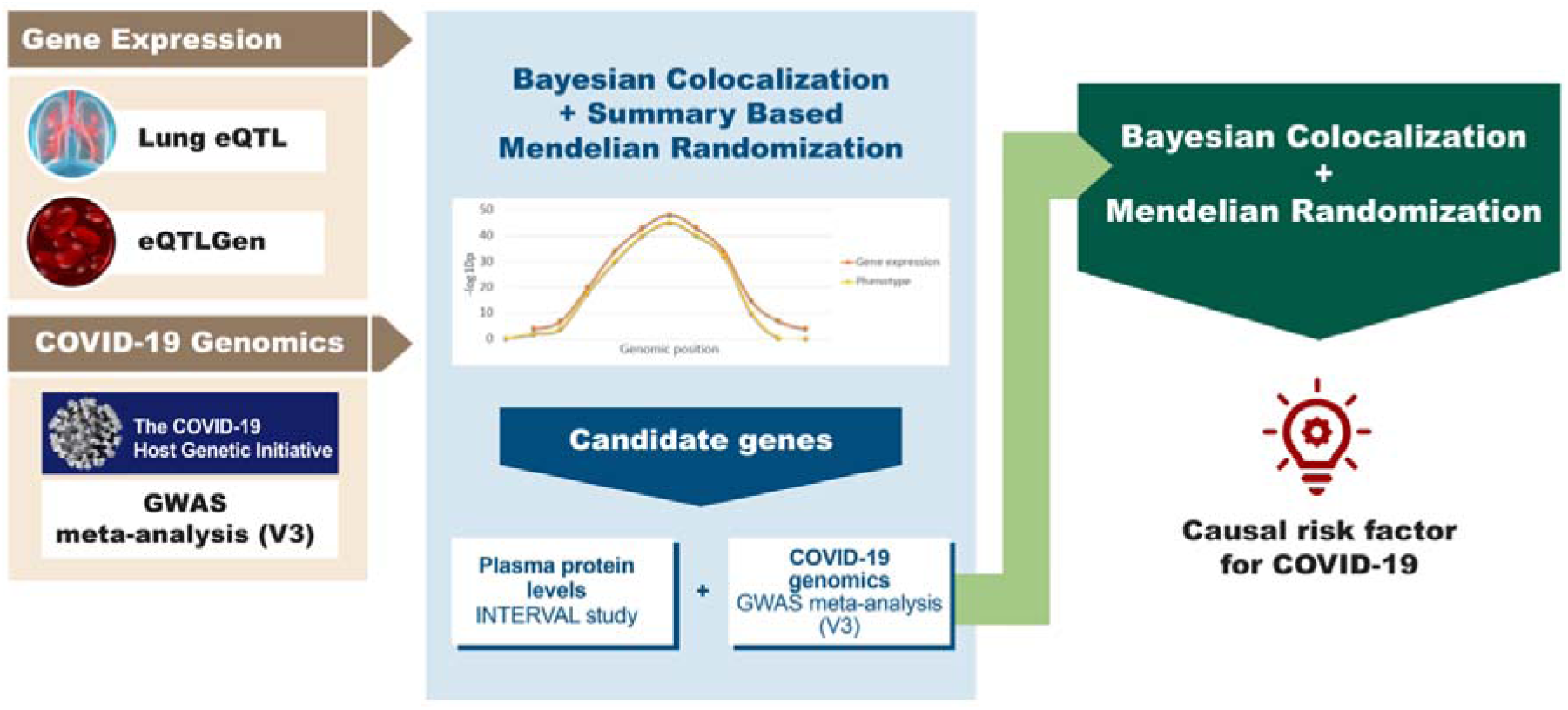
Study overview. The diagram summarizes the genomics datasets and analytic pipeline of the study. First, publicly available -omics datasets were obtained, which were later processed using integrative genomics (IG) methods (Bayesian Colocalization and Summary-based Mendelian Randomization) to identify potential candidate genes for COVID-19 phenotypes. Lastly, using a Bayesian Colocalization and Mendelian Randomization approach we explored the causal association between the plasma protein levels of the most promising candidate gene and COVID-19.

#### Lung gene expression

The expression of 14 unique genes in lung tissue co-localized (*PP*_*H4*_ being > 0.80) with severe COVID-19 associated loci (**Fig. 2a**) and 3 with the COVID-19 diagnosis associated loci (**Supplementary Table S1**). The majority of the gene associations identified in lung tissue were novel (i.e. not been identified by previous GWAS), and in most cases (10 out of 14), the genes met the suggestive statistical significance (*P*_*GWAS*_ < 5 × 10^−05^) rather than the genome-wide significance (*P*_*GWAS*_ < 5 × 10^−08^) (**Supplementary Table S1**). For example: *IL10RB* and *IFNAR2* (**Fig. 2a**) are interferon (IFN) receptor genes that co-localized with COVID-19 (*PP*_*H*4_ > 0.80); these genes were located in the same suggestive locus on chromosome 21 (sentinel SNP rs9976829, *P*_*GWAS*_ = 7.2 × 10^−07^ [COVID-19 hospitalization vs population] (The COVID-19 Host Genetics Initiative 2020)). In addition, results from the SMR show that the increased expression of *IFNAR2* in lung tissue was associated with decreased the risk of severe COVID-19 (**Supplementary Table S2**) and susceptibility to COVID-19 (**Fig. 2b**) (*P*_*SMR*_ < 0.001). SMR also identified a first-time association for the gene *OAS1* (**Fig. 2b and Supplementary Table S2**), which is an interferon stimulated gene involved in the cellular response to viral infection; increased expression of this gene was associated with decreased susceptibility to COVID-19.

**Fig. 2.**
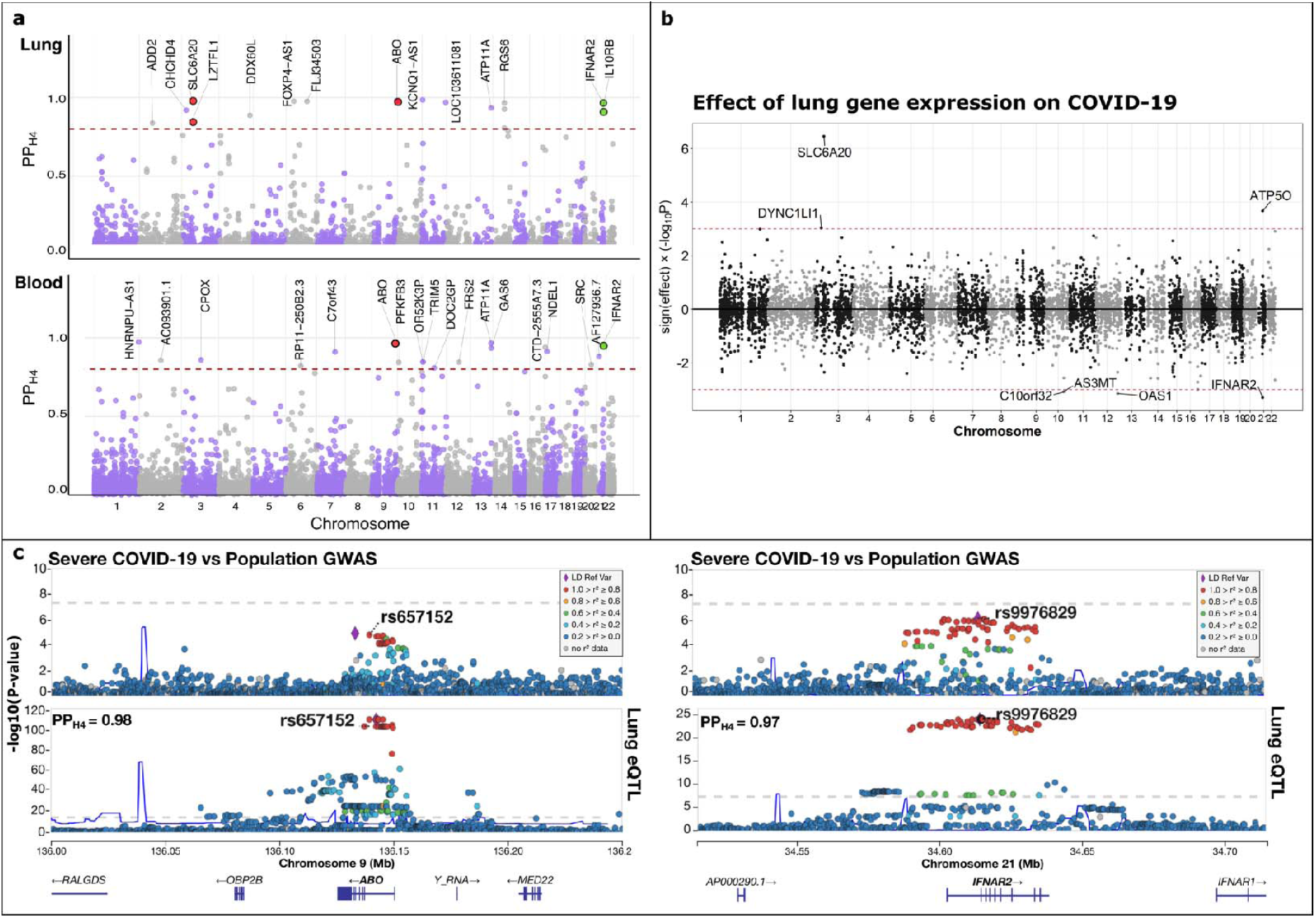
COVID-19 genomics and gene expression integration. a) Colocalization of COVID-19 (hospitalization vs population) with gene expression in lung and blood tissues, respectively. The circles represent the probability (y-axis) that a gene colocalizes (PP_H4_) with COVID-19 plotted against its chromosomal position (x-axis). The red dashed horizontal line represents the threshold of significance (PP_H4_ >0.80). Red and green circles highlight the genes within previously identified COVID-19 loci, and those involved in the interferon pathways, respectively. b) Results from the Summary Based Mendelian Randomization are displayed in this mirror Manhattan plot. The circles represent the association between susceptibility to COVID-19 (diagnosis vs population) and the gene expression (lung) multiplied by the direction of the effect (y-axis) plotted against the genes chromosomal position (x-axis). The dotted horizontal lines (red) represent the threshold of significance (P<0.001). SMR plot only shows the results that passed the heterogeneity test (see methods). c) Regional plots for severe COVID-19 (COVID-19 hospitalization vs population) and lung tissue cis-eQTL at the *ABO* (left panel) and *IFNAR2* (right panel) loci. Severe GWAS signals (top of the panel c) colocalize with the cis-QTL region (bottom of the panel C) at *ABO* (chromosome 9) and *IFNAR2* (chromosome 21). The circles represent the -log10 association p values (y-axis) of SNPs plotted against their chromosomal position (x-axis). The co-localized SNP is shown inside each plot and PP_H4_ is at the left top corner of the lung eQTL regional plots. Pairwise linkage disequilibrium (r^2^, calculated from the 1000 Genomes European population) in each region is colored with respect to the lead GWAS SNP within each region.

In addition to these novel gene associations, three co-localized genes (*LZTFL1, SLC6A20* and *ABO*) were within loci that have been previously associated with severe COVID-19 by GWAS(Ellinghaus et al. 2020) (**Fig. 2a**). SMR showed that increased *SLC6A20* expression in lung tissue was associated with increased the risk of severe COVID-19 (**Supplementary Table S2**) and susceptibility to COVID-19 (**Fig. 2b**). *ABO* gene expression co-localized with severe COVID-19 **Fig. 2c**. The colocalization between ABO gene expression and the COVID-19 susceptibility associated loci was not tested since the variants associated with this gene in the lung eQTL and eQTLGEN studies were not present in the COVID-19 HG meta-analysis for this phenotype.

#### Blood gene expression

In blood, the expression of 18 and 8 unique genes co-localized with COVID-19 severity (**Fig. 2a**) and susceptibility associated loci (**Supplementary Table S1**), respectively. The expression of *ABO* in blood co-localized with severe COVID-19 associated loci. In addition, *IFNAR2* expression in blood co-localized with both COVID-19 phenotypes, although its strongest colocalization (*PP*_*H4*_ = 0.96) was found with the COVID-19 hospitalization phenotype. Other first-time associations within COVID-19 suggestive loci included *HNRNPU*-*AS1, ATP11A* and *CTD*-*2555A7*.*3*. SMR identified 22 genes whose expression in blood was associated with COVID-19 phenotypes (**Supplementary Table S2**). Increased blood expression levels of *OAS1* and *CPOX* in blood were associated with decreased susceptibility to COVID-19 and increased risk of severe COVID-19, respectively.

### ABO plasma protein is a risk factor for COVID-19

The functional effects of genes are generally imparted through their translation into proteins. In order to strengthen the mechanistic association between the identified genes and COVID-19, we determined which of these genes were associated with both blood protein levels and COVID-19 phenotypes. We integrated the COVID-19 HG GWAS and blood protein GWAS from the INTERVAL study (which includes genome-wide associations between genetic variants and 3,622 blood proteins) using Coloc. Additionally, we applied MR to determined causal associations between protein and COVID-19.

Of the three protein-coding candidate genes (*ABO, IFNAR2*, and *ATP11A*) whose expression in both tissues co-localized with COVID-19 phenotypes, only *ABO* was present in the INTERVAL study. We found that ABO plasma protein levels co-localized with susceptibility to COVID-19 and its severity (**Fig. 3a and Supplementary Table S3**).

**Fig. 3.**
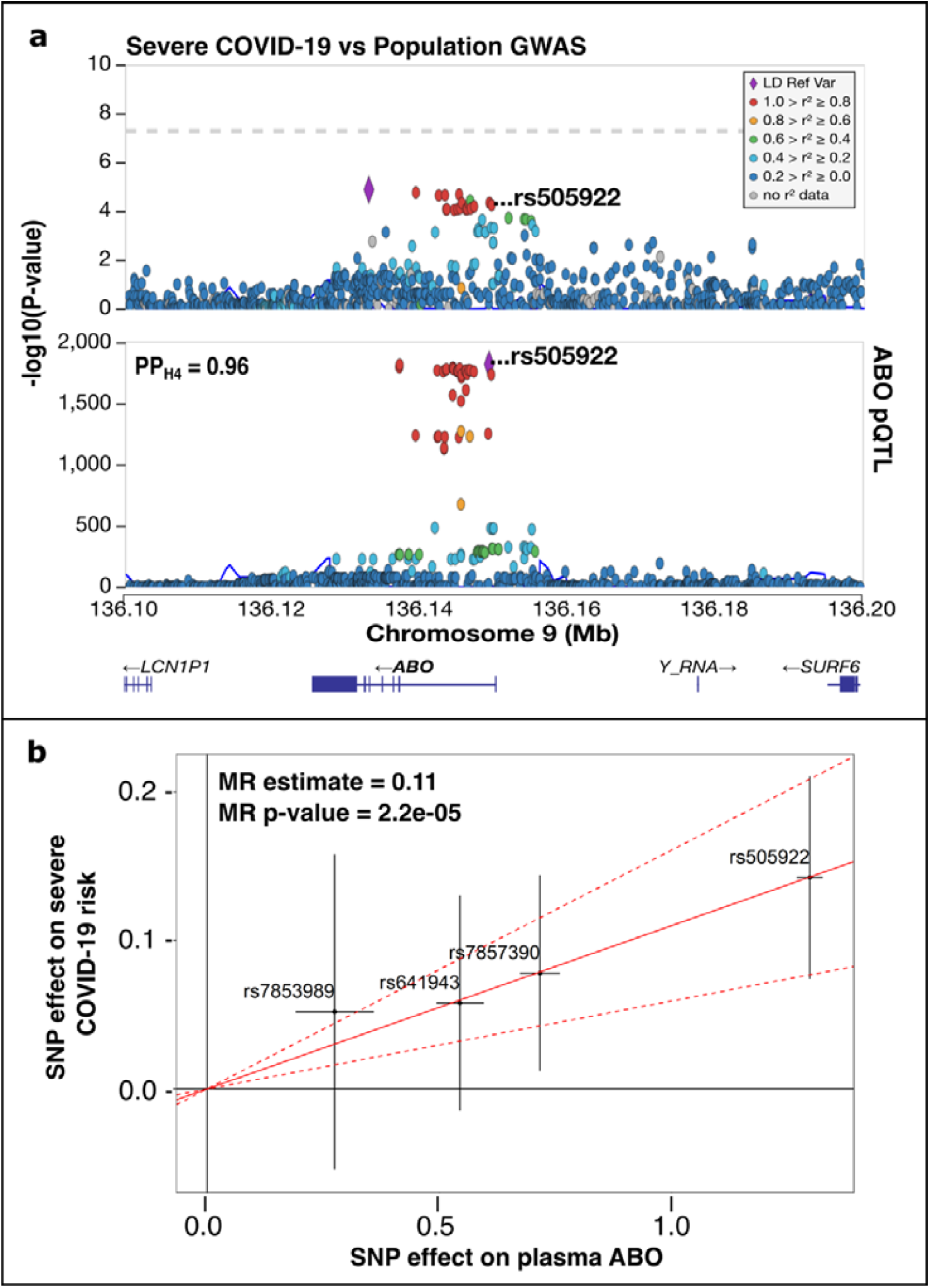
Bayesian Colocalization (a) and Mendelian Randomization (MR) (b) of ABO plasma protein and COVID-19. a) Regional plot describing severe COVID-19 GWAS (COVID-19 hospitalization vs population) (top of a) and plasma protein pQTL (bottom of A) at the *ABO* locus. The circles represent the -log10 association p values (y axis) of SNPs plotted against their chromosomal position (x-axis). The colocalized SNP is shown inside each plot and PP_H4_ is at the left top corner of the plasma protein pQTL regional plot. Pairwise linkage disequilibrium (r^2^, calculated from the 1000 Genomes European population) in each region is colored with respect to the lead GWAS SNP within each region. b) Inverse variance weighting (IVW) MR (IVW-MR) of ABO plasma proteins on risk of severe COVID-19 (COVID-19 hospitalization vs population). The x-axis represents the SNP effect on the plasma protein levels, and the y-axis the SNP effect on severe COVID-19. The variants used for the MR are shown inside the plot with error bars that represent the 95% confidence intervals. The slope of the solid red line is the instrumental variables regression estimate of the effect of the protein on severe COVID-19, with dashed red lines representing the 95 % confident interval. IVW-MR P-value and estimate are shown in the top left corner.

An IVW-MR test was conducted using four independent variants as instrumental variables (IVs) to investigate the causal relationship between ABO plasma protein and COVID-19. Based on the Cochran’s Q and MR-Egger intercept p-values the IV’s average effect on COVID-19 phenotypes did not demonstrate significant heterogeneity or horizontal pleiotropy (**Supplementary Table S4**). The results indicated a significant causal association between ABO plasma protein levels and COVID-19 phenotypes (*P* < 0.05) (**Fig. 3b and Supplementary Figure S1**). Each ABO-increasing allele increased the odds of COVID-19 by 7% and the odds of severe COVID-19 by 11% (**Fig. 3b**). In addition, we performed a sensitivity analysis to assess if the MR results were driven by a single SNP. **Supplementary Figures S2** and **S3** show that the exclusion of any of the IVs used for the MR did not significantly change the overall IVW-MR estimate.

## Discussion

Recent GWAS have revealed genetic loci that are significantly associated with COVID-19 (The COVID-19 Host Genetics Initiative 2020; Ellinghaus et al. 2020); however, since these loci often harbor multiple genes, the underlying genes or pathways responsible for COVID-19 remain unknown. To address this crucial gap in knowledge, we used integrative genomic methods, which unveiled several novel findings. First, we identified gene associations whose physiology plausibly relates to COVID-19. Second, we linked the effect of genetic variants to the gene expression of previously identified COVID-19 related-genes. Third, we showed that genetically-determined plasma ABO protein level is causally associated with the risk of severe COVID-19 and its susceptibility.

Although the chromosome 3p21 region has been previously associated with severe COVID-19, this locus encompasses six genes, making it hard to identify the precise gene responsible for the association with COVID-19 (Ellinghaus et al. 2020). We found that two genes within this locus (*SLC6A20 and LZTFL1*) co-localized with severe COVID-19. The *SLC6A20* protein product (sodium- and chloride-dependent transporter XTRP3) is involved in amino acid transport, with a role in the regulation of thymocyte selection (Simeoni et al. 2005) and negative regulation of T-cell activation (Arndt et al. 2011), suggesting its role in COVID-19 may be due altered immune responses. This protein may also interact with angiotensin converting enzyme 2 (ACE2), which is the putative SARS-CoV-2 receptor (Vuille-dit-Bille et al. 2015; Zhou et al. 2020b). Interestingly, both ACE2 and SLC6A20 gene and protein expression increase with age (Meier et al. 2018; Bunyavanich et al. 2020; Vuille-dit-Bille et al. 2020) with the lowest concentrations noted in children (small intestinal and nasal epitheliums); although children are susceptible to SARS-CoV-2 infection, they appear to be at very low risk of developing severe COVID-19 (Jordan et al. 2020). Consistent with these findings, another recent study has shown that a risk variant (rs11385942) for *SLC6A20* expression in human lung cells conferred an increased risk for severe COVID-19 (Ellinghaus et al. 2020, p. 19). *LZTFL1* encodes leucine zipper transcription factor-like protein 1, a protein involved in intracellular cargo trafficking that is linked to congenital ciliopathies; the mechanism of its association with COVID-19 is unclear.

Of the newly described gene associations with susceptibility to COVID-19 and severe COVID-19, *IL10RB, IFNAR2* and *OAS1* are notable given their role in the IFN pathways. The protein products of *IFNAR2* and *IL10RB* are components of the receptor complexes for type I and III IFNs, respectively, which are critical in the early host responses to a viral infection. The protein product of *OAS1* (2’-5’-oligoadenylate synthase 1), which is induced by IFNs, indirectly promotes viral RNA degradation and inhibition of viral replication. Our findings suggest that increased *IFNAR2* and *OAS1* gene expression decreases the susceptibility to COVID-19 and severe COVID-19. This is consistent with the concept of IFN dysregulation in severe COVID-19, in which patients mount a delayed and often blunted IFN response to the virus (Acharya et al. 2020; Made et al. 2020). Our findings support the results of a recent clinical trial showing that inhaled IFN-beta decreased the risk of severe COVID-19 (Balfour 2020).

Our results also show that a COVID-19 locus was associated with *ABO* gene expression in lung and blood tissues and plasma protein levels in blood. The *ABO* gene encodes for a protein responsible for the ABO blood groups. While the A and B allele carriers express glycosyltransferase activities that convert H antigen into A or B antigen, the O group protein lacks this enzymatic activity (due to a deletion in the gene [frameshift]). Previous studies have shown that blood type O individuals demonstrated a lower susceptibility to COVID-19 (Zhao et al. 2020; Zietz and Tatonetti 2020; Ellinghaus et al. 2020), and variants in the *ABO* have been associated with risk of severe COVID-19 (Ellinghaus et al. 2020). Using two-sample MR analysis, we showed that the ABO plasma protein is likely a causal risk factor for susceptibility to COVID-19 and severe COVID-19. The mechanism by which ABO protein modifies COVID-19 risk is unclear. In SARS-CoV a naturally occurring anti-A antibodies can inhibit spike protein-mediated cellular entry via the ACE2 receptor (Guillon et al. 2008) (this is also the putative entry mechanism for SARS-CoV-2), it has been speculated that this effect may also be found in SARS-CoV-2 (Zhao et al. 2020). Another possible explanation is that the A blood type is associated with an increased risk of cardiovascular disease (Wu et al. 2008), which is a known risk factors for severe COVID-19, while those with the O blood type are less likely to develop cardiovascular diseases. Furthermore, previous reports found an incidence of venous thromboembolism (VTE) of 27% in critically ill COVID-19 patients (Klok et al. 2020, p. 19). ABO blood type has been previously associated with risk of VTE (Wang et al. 2017); interestingly, the protein-increasing allele (C) of the top SNP (rs505922) used in the MR tests is also a risk variant for VTE (Trégouët et al. 2009). Further investigation is needed to understand the physiological role of ABO in the pathophysiology of COVID-19.

Our study had several limitations. Firstly, our analyses were limited by the number of genetic variants that overlapped between the COVID-19 HG meta-analysis and the datasets that were used for this study. Thus, we could not test some genes that may be of importance. Secondly, replication of our result in an independent dataset was not possible due to the lack of COVID-19 GWAS data available at the present time; therefore first-time associations identified by our analyses should be considered with caution. Lastly, the lung and blood -omics data used reflect the transcriptomic and proteomic profile under normal conditions, however it is likely that that these associations may change under stimulation by viral infection or acute inflammation.

## Conclusions

We used a multi-omics approach to identify several candidate genes that may be involved in the pathogenesis of COVID-19. The analyses presented here linked COVID-19 genomics to gene expression in lung and blood tissues. This approach revealed specific genes within previously implicated COVID-19 loci, and also identified new genes whose biology is consistent with COVID-19. Importantly, our analysis suggests that the ABO protein is a causal risk factor for severe COVID-19 and COVID-19 susceptibility.

## Data Availability

The results associated to this manuscript are available on the additional file. Specific datasets used to conduct this research are available as follow: The COVID-19 summary statistic can be accessed through the COVID-19 HG website (https://www.covid19hg.org/). Lung eQTL study expression and genotype data can be obtained through GEO platform accession number GSE23546 and the dbGaP Study Accession phs001745.v1.p1, respectively. Blood cis-eQTLs summary statistics can be accessed through the eQTLGen website (https://www.eqtlgen.org/cis-eqtls.html). The INTERVAL study pQTLs summary statistic are available through the European Genotype Archive (accession number EGAS00001002555).

## Declarations

### Funding

A.I.H.C. and S.M. are funded by MITACS Accelerate grant. D.D.S. holds the De Lazzari Family Chair at HLI and a Tier 1 Canada Research Chair in COPD. Y.B. holds a Canada Research Chair in Genomics of Heart and Lung Diseases.

### Competing interests

S.M. reports personal fees from Novartis and Boehringer-Ingelheim, outside the submitted work. W.T. reports fees to Institution from Roche-Ventana, AbbVie, Merck-Sharp-Dohme and Bristol-Myers-Squibb, outside the submitted work. M.v.d.B. reports research grants paid to Institution from Astra Zeneca, Novartis, outside the submitted work. D.D.S. reports research funding from AstraZeneca and received honoraria for speaking engagements from Boehringer Ingelheim and AstraZeneca over the past 36 months, outside of the submitted work.

### Ethics approval and consent to participate

Not applicable

### Consent to participate

Not applicable

### Consent for publication

Not applicable

### Code availability

Not applicable

### Authors’ contributions

DDS and AIHC designed the study. XL and AIHC conducted the data analyses. AIHC wrote the first draft of the manuscript. SM was involved in the interpretation of the results and critical review of the manuscript. CXY revised the manuscript. YB, PJ, WT, MB, DN, KH provided the lung cis-eQTLs dataset and revised the manuscript. DDS supervised this study, contributed to the interpretation of the results and revised the manuscript. All authors read and approved the final manuscript.

## Acknowledgments

The eQTL data from Laval University were generated from tissues obtained through the Quebec Research Respiratory Network Biobank, IUCPQ site. We acknowledge Compute Canada, which provided computational resources to conduct this research.

## Supplementary files

### Supplementary Tables

Table S1: Genes associated to COVID-19 through colocalization.

Table S2: Genes associated to COVID-19 through Summary-based Mendelian Randomization.

Table S3. COVID-19 phenotypes and ABO plasma protein colocalization.

Table S4. Inverse variance weighting mendelian randomization (IVW-MR) of ABO plasma protein and COVID-19.

### Supplementary Figures

Figure S1. Mendelian Randomization of ABO plasma protein and susceptibility to COVID-19.

Figure S2. Mendelian Randomization sensitivity analysis for dependence on individual instruments (severe COVID-19 and ABO plasma protein).

Figure S3. Mendelian Randomization (MR) sensitivity analysis for dependence on individual instruments (susceptibility of COVID-19 and ABO plasma protein).

## Notes

### Author Declarations

Not applicable. Publicly available third-party data sets were used for this.

## References

1000 Genomes Project Consortium, Auton A, Brooks LD, et al (2015) A global reference for human genetic variation. Nature 526:68–74. https://doi.org/10.1038/nature15393

Acharya D, Liu G, Gack MU (2020) Dysregulation of type I interferon responses in COVID-19. Nature Reviews Immunology 20:397–398. https://doi.org/10.1038/s41577-020-0346-x

Arndt B, Krieger T, Kalinski T, et al (2011) The transmembrane adaptor protein SIT inhibits TCR-mediated signaling. PLoS ONE 6:e23761. https://doi.org/10.1371/journal.pone.0023761

Balfour H (2020) Inhaled interferon beta therapy shows promise in COVID-19 trial. In: European Pharmaceutical Review. https://www.europeanpharmaceuticalreview.com/news/123994/inhaled-interferon-beta-therapy-shows-promise-in-covid-19-trial/. Accessed 7 Aug 2020

Bowden J, Davey Smith G, Burgess S (2015) Mendelian randomization with invalid instruments: effect estimation and bias detection through Egger regression. Int J Epidemiol 44:512–525. https://doi.org/10.1093/ije/dyv080

Bunyavanich S, Do A, Vicencio A (2020) Nasal Gene Expression of Angiotensin-Converting Enzyme 2 in Children and Adults. JAMA 323:2427–2429. https://doi.org/10.1001/jama.2020.8707

Bycroft C, Freeman C, Petkova D, et al (2018) The UK Biobank resource with deep phenotyping and genomic data. Nature 562:203–209. https://doi.org/10.1038/s41586-018-0579-z

Cano-Gamez E, Trynka G (2020) From GWAS to Function: Using Functional Genomics to Identify the Mechanisms Underlying Complex Diseases. Front Genet 11:. https://doi.org/10.3389/fgene.2020.00424

Chang CC, Chow CC, Tellier LC, et al (2015) Second-generation PLINK: rising to the challenge of larger and richer datasets. Gigascience 4:. https://doi.org/10.1186/s13742-015-0047-8

Di Angelantonio E, Thompson SG, Kaptoge S, et al (2017) Efficiency and safety of varying the frequency of whole blood donation (INTERVAL): a randomised trial of 45L000 donors. Lancet 390:2360–2371. https://doi.org/10.1016/S0140-6736(17)31928-1

Dong E, Du H, Gardner L (2020) An interactive web-based dashboard to track COVID-19 in real time. Lancet Infect Dis 20:533–534. https://doi.org/10.1016/S1473-3099(20)30120-1

Ellinghaus D, Degenhardt F, Bujanda L, et al (2020) Genomewide Association Study of Severe Covid-19 with Respiratory Failure. N Engl J Med. https://doi.org/10.1056/NEJMoa2020283

Giambartolomei C, Vukcevic D, Schadt EE, et al (2014) Bayesian Test for Colocalisation between Pairs of Genetic Association Studies Using Summary Statistics. PLOS Genetics 10:e1004383. https://doi.org/10.1371/journal.pgen.1004383

Guillon P, Clément M, Sébille V, et al (2008) Inhibition of the interaction between the SARS-CoV Spike protein and its cellular receptor by anti-histo-blood group antibodies. Glycobiology 18:1085–1093. https://doi.org/10.1093/glycob/cwn093

Hao K, Bossé Y, Nickle DC, et al (2012) Lung eQTLs to help reveal the molecular underpinnings of asthma. PLoS Genet 8:e1003029. https://doi.org/10.1371/journal.pgen.1003029

Jordan RE, Adab P, Cheng KK (2020) Covid-19: risk factors for severe disease and death. BMJ 368:. https://doi.org/10.1136/bmj.m1198

Klok FA, Kruip MJHA, van der Meer NJM, et al (2020) Incidence of thrombotic complications in critically ill ICU patients with COVID-19. Thromb Res 191:145–147. https://doi.org/10.1016/j.thromres.2020.04.013

Made CI van der, Simons A, Schuurs-Hoeijmakers J, et al (2020) Presence of Genetic Variants Among Young Men With Severe COVID-19. JAMA. https://doi.org/10.1001/jama.2020.13719

Meier C, Camargo SM, Hunziker S, et al (2018) Intestinal IMINO transporter SIT1 is not expressed in human newborns. American Journal of Physiology-Gastrointestinal and Liver Physiology 315:G887–G895. https://doi.org/10.1152/ajpgi.00318.2017

Simeoni L, Posevitz V, Kölsch U, et al (2005) The transmembrane adapter protein SIT regulates thymic development and peripheral T-cell functions. Mol Cell Biol 25:7557–7568. https://doi.org/10.1128/MCB.25.17.7557-7568.2005

Staley OYJ (2020) MendelianRandomization: Mendelian Randomization Package. Version 0.4.3URL https://CRAN.R-project.org/package=MendelianRandomization

Sun BB, Maranville JC, Peters JE, et al (2018) Genomic atlas of the human plasma proteome. Nature 558:73–79. https://doi.org/10.1038/s41586-018-0175-2

Swerdlow DI, Kuchenbaecker KB, Shah S, et al (2016) Selecting instruments for Mendelian randomization in the wake of genome-wide association studies. Int J Epidemiol 45:1600–1616. https://doi.org/10.1093/ije/dyw088

The COVID-19 Host Genetics Initiative (2020) The COVID-19 Host Genetics Initiative, a global initiative to elucidate the role of host genetic factors in susceptibility and severity of the SARS-CoV-2 virus pandemic. European Journal of Human Genetics 28:715–718. https://doi.org/10.1038/s41431-020-0636-6

Trégouët D-A, Heath S, Saut N, et al (2009) Common susceptibility alleles are unlikely to contribute as strongly as the FV and ABO loci to VTE risk: results from a GWAS approach. Blood 113:5298–5303. https://doi.org/10.1182/blood-2008-11-190389

Võsa U, Claringbould A, Westra H-J, et al (2018) Unraveling the polygenic architecture of complex traits using blood eQTL metaanalysis. bioRxiv 447367. https://doi.org/10.1101/447367

Vuille-dit-Bille RN, Camargo SM, Emmenegger L, et al (2015) Human intestine luminal ACE2 and amino acid transporter expression increased by ACE-inhibitors. Amino Acids 47:693–705. https://doi.org/10.1007/s00726-014-1889-6

Vuille-dit-Bille RN, Liechty KW, Verrey F, Guglielmetti LC (2020) SARS-CoV-2 receptor ACE2 gene expression in small intestine correlates with age. Amino Acids. https://doi.org/10.1007/s00726-020-02870-z

Wang Z, Dou M, Du X, et al (2017) Influences of ABO blood group, age and gender on plasma coagulation factor VIII, fibrinogen, von Willebrand factor and ADAMTS13 levels in a Chinese population. PeerJ 5:. https://doi.org/10.7717/peerj.3156

Westra H-J, Peters MJ, Esko T, et al (2013) Systematic identification of trans eQTLs as putative drivers of known disease associations. Nature Genetics 45:1238–1243. https://doi.org/10.1038/ng.2756

Wu O, Bayoumi N, Vickers MA, Clark P (2008) ABO(H) blood groups and vascular disease: a systematic review and meta-analysis. J Thromb Haemost 6:62–69. https://doi.org/10.1111/j.1538-7836.2007.02818.x

Yang J, Lee SH, Goddard ME, Visscher PM (2011) GCTA: a tool for genome-wide complex trait analysis. Am J Hum Genet 88:76–82. https://doi.org/10.1016/j.ajhg.2010.11.011

Zhao J, Yang Y, Huang H, et al (2020) Relationship between the ABO Blood Group and the COVID-19 Susceptibility. medRxiv 2020.03.11.20031096. https://doi.org/10.1101/2020.03.11.20031096

Zhou F, Yu T, Du R, et al (2020a) Clinical course and risk factors for mortality of adult inpatients with COVID-19 in Wuhan, China: a retrospective cohort study. The Lancet 395:1054–1062. https://doi.org/10.1016/S0140-6736(20)30566-3

Zhou P, Yang X-L, Wang X-G, et al (2020b) A pneumonia outbreak associated with a new coronavirus of probable bat origin. Nature 579:270–273. https://doi.org/10.1038/s41586-020-2012-7

Zhu N, Zhang D, Wang W, et al (2020) A Novel Coronavirus from Patients with Pneumonia in China, 2019. New England Journal of Medicine 382:727–733. https://doi.org/10.1056/NEJMoa2001017

Zhu Z, Zhang F, Hu H, et al (2016) Integration of summary data from GWAS and eQTL studies predicts complex trait gene targets. Nat Genet 48:481–487. https://doi.org/10.1038/ng.3538

Zietz M, Tatonetti NP (2020) Testing the association between blood type and COVID-19 infection, intubation, and death. medRxiv. https://doi.org/10.1101/2020.04.08.20058073

