## Supplementary Figures for "Multi-omics highlights ABO plasma protein as a causal risk factor for COVID-19"

*Human Genetics*

Ana I. Hernández Cordero^1^, Xuan Li^1^, Stephen Milne^1,2,3^, Chen Xi Yang^1^, Yohan Bossé^4^, Philippe Joubert^4^, Wim Timens^5^, Maarten van den Berge^6^, David Nickle^7^, Ke Hao^8^, Don D. Sin^1,2^

^1^Centre for Heart Lung Innovation, University of British Columbia, Vancouver, BC, Canada

^2^ Division of Respiratory Medicine, Faculty of Medicine, University of British Columbia, Vancouver, BC, Canada

^3^ Faculty of Medicine and Health, University of Sydney, Sydney, New South Wales, Australia

^4^ Institut universitaire de cardiologie et de pneumologie de Québec - Université Laval, Québec City, QC, Canada

^5^ University of Groningen, University Medical Centre Groningen, Department of Pathology and Medical Biology, Groningen, The Netherlands

^6^ University of Groningen, University Medical Center Groningen, Department of Pulmonary Diseases, Groningen, The Netherlands

^7^ Merck Research Laboratories, Genetics and Pharmacogenomics, Boston, Massachusetts, United States of America

^8^ Department of Genetics and Genomic Sciences and Icahn Institute for Data Science and Genomic Technology, Icahn School of Medicine at Mount Sinai, New York, NY, United States of America

**Corresponding author:** Ana I. Hernández Cordero, PhD

**Address:** Centre for Heart Lung Innovation, Room 164,

1081 Burrard Street, St. Paul's Hospital, Vancouver, BC V6Z 1Y6

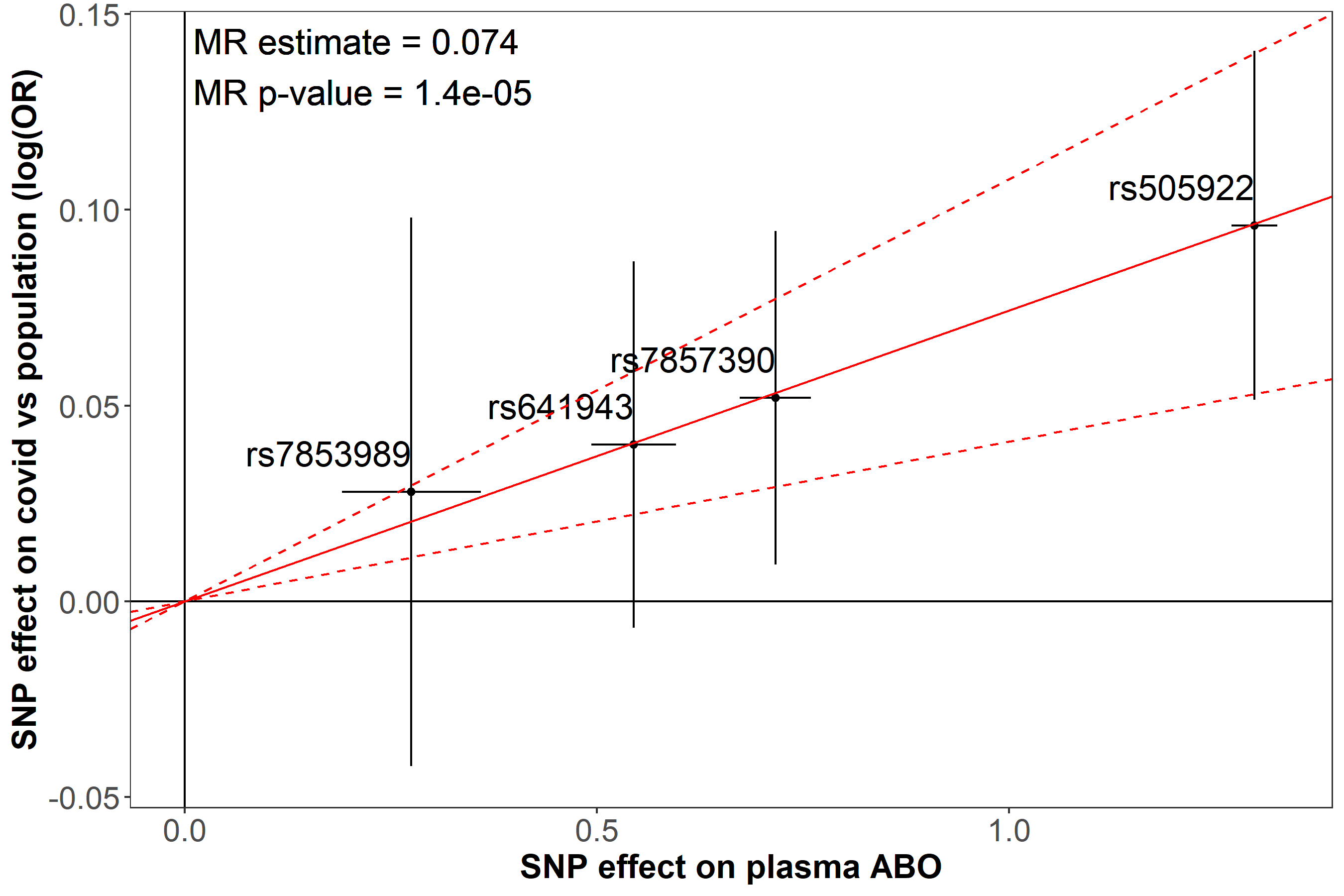

**Supplementary Figure S1. Mendelian Randomization of ABO plasma protein and susceptibility to COVID-19.** Susceptibility to COVID-19 defined as a positive COVID-19 diagnosis versus the general population.

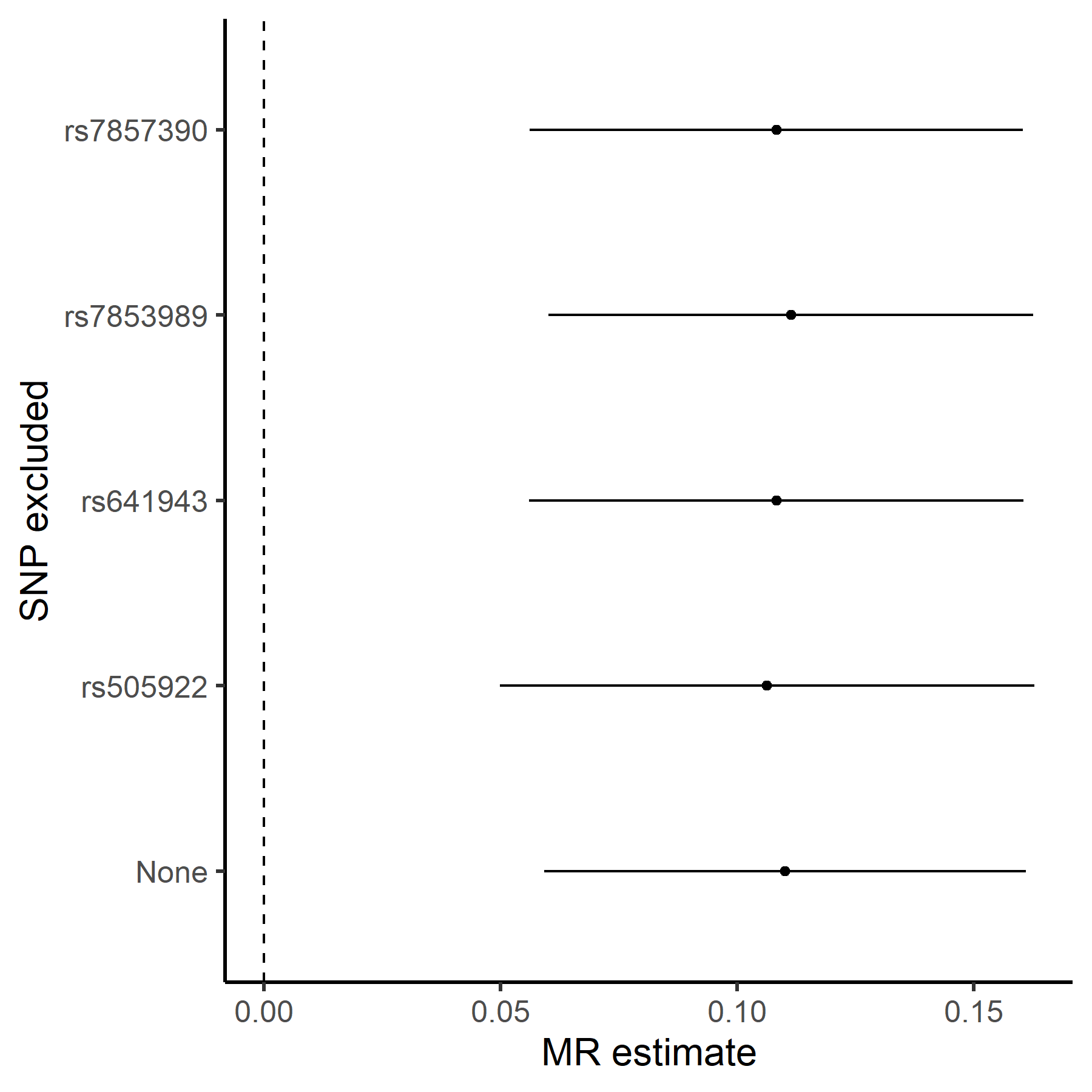

**Supplementary Figure S2. Mendelian Randomization sensitivity analysis for dependence on individual instruments (severe COVID-19 and ABO plasma protein).** The dots represented the MR per allele effect (x-axis) estimated from of each inverse variance weighting (IVW) MR (IVW-MR) re-run (y-axis) with the 95% confidence interval. The SNPs showed in the y-axis represent the excluded SNPs form each of the 4 re-runs, while ‘none’ represent the IVW-MR analysis with the four independent SNP. The overall MR estimate remained significant despite the systematic exclusion of each SNP, indicating that the overall estimate was not dependent on the effects of a single instrumental variable.

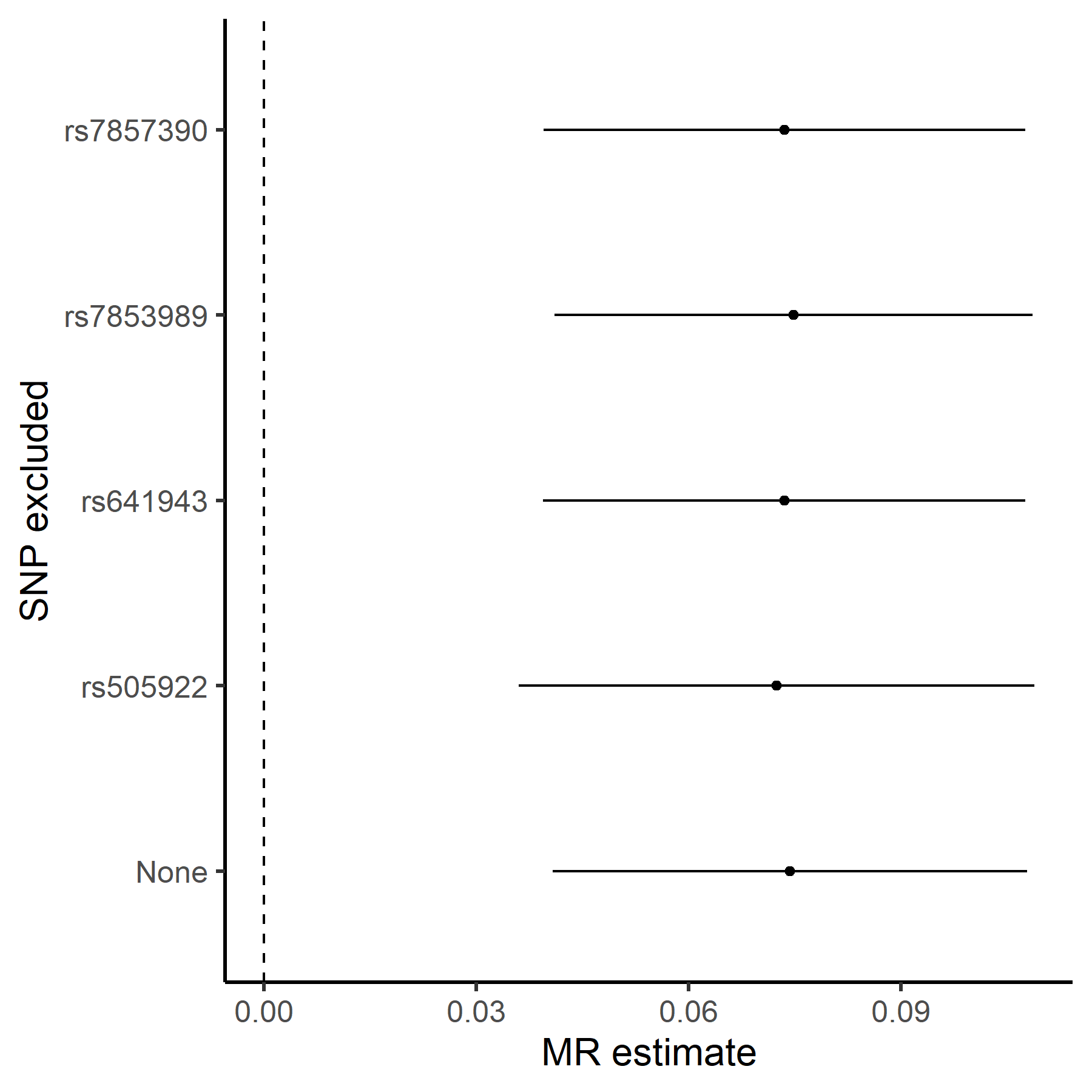

**Supplementary Figure S3. Mendelian Randomization (MR) sensitivity analysis for dependence on individual instruments (susceptibility of COVID-19 and ABO plasma protein).** The dots represented the MR per allele effect (x-axis) estimated from of each inverse variance weighting (IVW) MR (IVW-MR) re-run (y-axis) with the 95% confidence interval. The SNPs showed in the y-axis represent the excluded SNPs form each of the 4 re-runs, while ‘none’ represent the IVW-MR analysis with the four independent SNP. The overall MR estimate remained significant despite the systematic exclusion of each SNP, indicating that the overall estimate was not dependent on the effects of a single instrumental variable.
