## Supplementary Tables for "Multi-omics highlights ABO plasma protein as a causal risk factor for COVID-19"

*Human Genetics*

Ana I. Hernández Cordero^1^, Xuan Li^1^, Stephen Milne^1,2,3^, Chen Xi Yang^1^, Yohan Bossé^4^, Philippe Joubert^4^, Wim Timens^5^, Maarten van den Berge^6^, David Nickle^7^, Ke Hao^8^, Don D. Sin^1,2^

^1^Centre for Heart Lung Innovation, University of British Columbia, Vancouver, BC, Canada

^2^ Division of Respiratory Medicine, Faculty of Medicine, University of British Columbia, Vancouver, BC, Canada

^3^ Faculty of Medicine and Health, University of Sydney, Sydney, New South Wales, Australia

^4^ Institut universitaire de cardiologie et de pneumologie de Québec - Université Laval, Québec City, QC, Canada

^5^ University of Groningen, University Medical Centre Groningen, Department of Pathology and Medical Biology, Groningen, The Netherlands

^6^ University of Groningen, University Medical Center Groningen, Department of Pulmonary Diseases, Groningen, The Netherlands

^7^ Merck Research Laboratories, Genetics and Pharmacogenomics, Boston, Massachusetts, United States of America

^8^ Department of Genetics and Genomic Sciences and Icahn Institute for Data Science and Genomic Technology, Icahn School of Medicine at Mount Sinai, New York, NY, United States of America

**Corresponding author:** Ana I. Hernández Cordero, PhD

**Address:** Centre for Heart Lung Innovation, Room 164,

1081 Burrard Street, St. Paul's Hospital, Vancouver, BC V6Z 1Y6

**Supplementary Table S1: Genes associated to COVID-19 through colocalization.**

| Dataset | COVID-19 phenotype**^‡^** | Gene Symbol | Location^†^ | PP.H4 |
| --- | --- | --- | --- | --- |
| Lung eQTL (Lung tissue) | Susceptibility to COVID-19 | *WNK1* | 12p13.33 | 0.858 |
|  |  | *DYNC1LI1* | 3p22.3 | 0.839 |
|  |  | *IFNAR2* | 21q22.11 | 0.818 |
|  | Severe COVID-19 | ***SLC6A20*** | 3p21.31 | 0.987 |
|  |  | *KCNQ1-AS1* | 11p15.4 | 0.985 |
|  |  | ***ABO*** | 9q34.2 | 0.980 |
|  |  | *FOXP4-AS1* |  | 0.977 |
|  |  | *FLJ34503* |  | 0.976 |
|  |  | *IFNAR2* | 21q22.11 | 0.972 |
|  |  | *LOC103611081* |  | 0.970 |
|  |  | *ATP11A* | 13q34 | 0.935 |
|  |  | *RGS6* | 14q24.2 | 0.927 |
|  |  | *CHCHD4* | 3p25.1 | 0.920 |
|  |  | *IL10RB* | 21q22.11 | 0.917 |
|  |  | *DDX60L* | 4q32.3 | 0.887 |
|  |  | ***LZTFL1*** | 3p21.31 | 0.852 |
|  |  | *ADD2* | 2p13.3 | 0.837 |
| eQTLGEN  (Blood tissue) | Susceptibility to COVID-19 | *RNF24* | 20p13 | 0.968 |
|  |  | *ITGAD* | 16p11.2 | 0.949 |
|  |  | *NIPAL2* | 8q22.2 | 0.914 |
|  |  | *DCUN1D1* | 3q27.1 | 0.878 |
|  |  | *SRC* | 20q11.23 | 0.873 |
|  |  | *PLA2G15* | 16q22.1 | 0.868 |
|  |  | *IFNAR2* | 21q22.11 | 0.839 |
|  |  | *RP5-1085F17.3* | 20q11.21 | 0.804 |
|  | Severe COVID-19 | *HNRNPU-AS1* | 1q44 | 0.974 |
|  |  | ***ABO*** | 9q34.2 | 0.971 |
|  |  | *ATP11A* | 13q34 | 0.966 |
|  |  | *IFNAR2* | 21q22.11 | 0.957 |
|  |  | *CTD-2555A7.3* | 16q24.3 | 0.939 |
|  |  | *GAS6* | 13q34 | 0.935 |
|  |  | *NDEL1* | 17p13.1 | 0.913 |
|  |  | *C7orf43* | 7q22.1 | 0.910 |
|  |  | *AF127936.7* | 21q11.2 | 0.880 |
|  |  | *CPOX* | 3q12.1 | 0.860 |
|  |  | *AC093901.1* | 2q14.2 | 0.857 |
|  |  | *OR52K3P* | 11p15.4 | 0.847 |
|  |  | *TRIM5* | 11p15.4 | 0.846 |
|  |  | *FRS2* | 12q15 | 0.845 |
|  |  | *PFKFB3* | 10p15.1 | 0.844 |
|  |  | *SRC* | 20q11.23 | 0.830 |
|  |  | *RP11-250B2.3* | 6q14.1 | 0.821 |
|  |  | *DOC2GP* | 11q13.2 | 0.808 |

^†^Cytogenetic location. **^‡^** Susceptibility to COVID-19 defined as a positive COVID-19 diagnosis versus the general population and severe COVID-19 defined as hospitalization for COVID-19 versus the general population. Bolded genes’ symbol represents those genes in genome-significant COVID-19 loci (P_GWAS_ < 5 × 10^-08^). Underlined genes’ symbol represents those in suggestive COVID-19 loci (P_GWAS_ < 5 × 10^-05^).

**Supplementary Table S2: Genes associated to COVID-19 through Summary-based Mendelian Randomization.**

| Dataset | COVID-19 phenotype**^‡^** | Gene Symbol | Location^†^ | beta | se | P_SMR_ | P_HEIDI_ |
| --- | --- | --- | --- | --- | --- | --- | --- |
| Lung eQTL  (Lung tissue) | Susceptibility to COVID-19 | ***SLC6A20*** | 3p21.31 | 0.28 | 0.06 | 3.50 × 10^-07^ | 0.36 |
|  |  | *ATP5O* | 21q22.11 | 1.44 | 0.39 | 2.13 × 10^-04^ | 0.45 |
|  |  | *IFNAR2* | 21q22.11 | -0.65 | 0.19 | 4.95 × 10^-04^ | 0.37 |
|  |  | *OAS1* | 12q24.13 | -0.14 | 0.04 | 7.07 × 10^-04^ | 0.06 |
|  |  | *C10orf32* | 10q24.32 | -0.33 | 0.10 | 8.14 × 10^-04^ | 0.47 |
|  |  | *AS3MT* | 10q24.32 | -0.25 | 0.08 | 8.48 × 10^-04^ | 0.10 |
|  |  | *DYNC1LI1* | 3p22.3 | 1.17 | 0.35 | 9.44 × 10^-04^ | 0.39 |
|  | Severe COVID-19 | ***SLC6A20*** | 3p21.31 | 0.58 | 0.09 | 1.33 × 10^-10^ | 0.41 |
|  |  | *IFNAR2* | 21q22.11 | -1.33 | 0.30 | 1.04 × 10^-05^ | 0.13 |
| eQTLGEN  (Blood tissue) | Susceptibility to COVID-19 | *RNF24* | 20p13 | 0.42 | 0.11 | 8.52 × 10^-05^ | 0.73 |
|  |  | *ITGAD* | 16p11.2 | 0.29 | 0.07 | 9.69 × 10^-05^ | 0.15 |
|  |  | *NIPAL2* | 8q22.2 | -0.19 | 0.05 | 2.96 × 10^-04^ | 0.16 |
|  |  | *COX20* | 1q44 | 0.27 | 0.08 | 3.72 × 10^-04^ | 0.11 |
|  |  | *RP5-1085F17.3* | 20q11.21 | 0.21 | 0.06 | 5.15 × 10^-04^ | 0.61 |
|  |  | *OAS1* | 12q24.13 | -0.12 | 0.03 | 6.42 × 10^-04^ | 0.16 |
|  |  | *C10orf32* | 10q24.32 | -0.12 | 0.04 | 6.64 × 10^-04^ | 0.63 |
|  |  | *ESPNL* | 2q37.3 | 0.51 | 0.15 | 7.00 × 10^-04^ | 0.48 |
|  |  | *EXOSC6* | 16q22.1 | -0.12 | 0.04 | 9.47 × 10^-04^ | 0.70 |
|  |  | *ATP8B4* | 15q21.2 | -0.16 | 0.05 | 9.95 × 10^-04^ | 0.26 |
|  | Severe COVID-19 | *CTD-2555A7.3* | 16q24.3 | 0.51 | 0.14 | 2.39 × 10^-04^ | 0.65 |
|  |  | *GAS6* | 13q34 | -0.74 | 0.20 | 2.43 × 10^-04^ | 0.56 |
|  |  | *ATP11A* | 13q34 | -1.76 | 0.49 | 2.95 × 10^-04^ | 0.08 |
|  |  | *NDEL1* | 17p13.1 | -0.92 | 0.26 | 3.63 × 10^-04^ | 0.37 |
|  |  | *CEP120* | 5q23.2 | 1.29 | 0.36 | 4.09 × 10^-04^ | 0.47 |
|  |  | *CPOX* | 3q12.1 | -0.74 | 0.21 | 4.23 × 10^-04^ | 0.25 |
|  |  | *OR52K3P* | 11p15.4 | 0.14 | 0.04 | 4.51 × 10^-04^ | 0.58 |
|  |  | *AC093901.1* | 2q14.2 | 0.66 | 0.19 | 4.65 × 10^-04^ | 0.51 |
|  |  | *TRIM5* | 11p15.4 | -0.34 | 0.10 | 6.42 × 10^-04^ | 0.32 |
|  |  | *DOC2GP* | 11q13.2 | -0.13 | 0.04 | 6.55 × 10^-04^ | 0.16 |
|  |  | *OR52K2* | 11p15.4 | 0.39 | 0.12 | 8.58 × 10^-04^ | 0.40 |
|  |  | *LAYN* | 11q23.1 | -1.25 | 0.38 | 9.98 × 10^-04^ | 0.36 |

^†^Cytogenetic location. **^‡^** Susceptibility to COVID-19 defined as a positive COVID-19 diagnosis versus the general population and severe COVID-19 defined as hospitalization for COVID-19 versus the general population. Bolded genes’ symbols represent those genes in genome-significant COVID-19 loci (P_GWAS_ < 5 × 10^-08^). Underlined genes’ symbols represent those in suggestive COVID-19 loci (P_GWAS_ < 5 × 10^-05^).

**Supplementary Table S3. COVID-19 phenotypes and ABO plasma protein colocalization.**

| COVID-19  phenotype | Protein | Location^†^ | rsID | Allele (Alt/Ref) | pQTL effect^‡^ | GWAS effect (P_GWAS_) | PP_H4_ |
| --- | --- | --- | --- | --- | --- | --- | --- |
| Susceptibility to COVID-19 | ABO | 9q34.2 | rs505922 | T/C | -1.3 | -0.096 (2.4 × 10^-05^) | 0.97 |
| Severe COVID-19 |  |  |  |  |  | -0.14 (4.4 × 10^-05^) | 0.96 |

^†^Cytogenetic location. ^‡^No nominal P could be obtained for the ABO protein quantitative trait locus (pQTL) because of the high significance of the locus (close to 0). Susceptibility to COVID-19 defined as a positive COVID-19 diagnosis versus the general population and severe COVID-19 defined as hospitalization for COVID-19 versus the general population. ‘Alt’ allele is the effect allele. ‘GWAS effect’ refers to the allelic effect estimated in the COVID-19 association analyses.

Supplementary Table S4. Inverse variance weighting mendelian randomization (IVW-MR) of ABO plasma protein and COVID-19.

| COVID-19 phenotype^†^ | IVW-MR est | IVW-MR SE | IVW-MR P | P_Cochran’s Q test_ | P_Egger Intercept_ |
| --- | --- | --- | --- | --- | --- |
| Susceptibility to COVID-19 | 0.07 | 0.02 | 1.39 × 10^-05^ | 0.99 | 0.97 |
| Severe COVID-19 | 0.11 | 0.03 | 2.21 × 10^-05^ | 0.96 | 0.93 |

**^†^** Susceptibility to COVID-19 defined as a positive COVID-19 diagnosis versus the general population and severe COVID-19 defined as hospitalization for COVID-19 versus the general population.
